# Clinical Stability under FF/UMEC/VI Triple Inhaled Therapy: A 12-Month Real Life Retrospective Observational Study

**DOI:** 10.1101/2025.10.20.25338377

**Authors:** Mauro Maniscalco, Claudio Candia, Francesco Pennisi, Alfio Pennisi, Giuseppe De Simone, Pasquale Ambrosino

## Abstract

Chronic Obstructive Pulmonary Disease (COPD) remains a leading cause of morbidity and mortality worldwide. For patients with a high disease burden, triple therapy with fluticasone furoate/umeclidinium bromide/vilanterol trifenatate (FF/UMEC/VI) has demonstrated significant benefits in health-related quality of life (HRQoL), exacerbation reduction, and lung function improvement. Efforts have been made to define clinical stability (CS), yet real-world data on CS during FF/UMEC/VI therapy remain limited.

This retrospective study aimed to assess the prevalence of CS after 12 months (T12) of FF/UMEC/VI treatment in COPD patients. CS was defined as the concurrent presence at T12 of: no acute exacerbations in the prior 12 months, a ≥ 2-point improvement in COPD Assessment Test (CAT) score from baseline, and a forced expiratory volume in one second (FEV□) decline < 100 mL.

A total of 47 patients was included. Of them, 10 (21.3%) achieved CS. These individuals had a lower baseline exacerbation rate (p = 0.020) and a trend toward better baseline lung function. They also demonstrated greater improvement in six-minute walking distance compared with non-CS patients (P = 0.048).

These findings suggest that CS is attainable in routine clinical practice, with prevalence comparable to that observed in clinical trials. Patients achieving CS tended to have milder disease, indicating potential benefits of earlier FF/UMEC/VI initiation. Further multicenter prospective studies are warranted to validate CS as a meaningful outcome in pulmonary rehabilitation and to identify predictors of treatment success.

## 1. Introduction

Chronic Obstructive Pulmonary Disease (COPD) is a leading cause of morbidity and mortality worldwide [1]. It is characterized by fixed airway obstruction, limitations in daily activities, and frequent acute exacerbations [2]. On one hand, COPD significantly burdens patients by severely impacting their health-related quality of life (HRQoL) [3]; on the other hand, it imposes substantial costs on healthcare systems [4]. The majority of COPD-related expenditures stem from acute exacerbations and comorbidities, particularly cardiovascular diseases (CVD), which are more prevalent in this patient population [5].

The introduction of triple inhaled therapy with Fluticasone Furoate/Umeclidinium Bromide/Vilanterol Trifenatate (FF/UMEC/VI) has had a significant impact on key outcomes, with reported improvements in HRQoL, exacerbation frequency, and lung function, as shown by meta-analytical data [6]. Recently, the concept of clinical stability (CS) in COPD has been introduced through post-hoc analyses of major multicenter, placebo-controlled clinical trials. Among the various definitions proposed, a promising one is that of Beeh *et al*. [7], which encompasses the three most relevant aspects of COPD burden. In their recent publication [7], the authors described a three-component definition of CS, which includes: absence of acute exacerbations, a reduction in the COPD Assessment Test (CAT) score of at least 2 points, and a decline in forced expiratory volume in one second (FEV_1_) of no more than 100 mL/year.

Given the lack of real-world data on clinical stability and the potential value of this novel endpoint, the aim of the present study is to retrospectively evaluate the prevalence of clinical stability, as defined by Beeh *et al*. [7], in a cohort of patients with COPD.

## 2. Patients and Methods

### 2.1 Study Design

This study was designed as a retrospective, single-center observational protocol. Medical records of consecutive patients admitted to our Institute for a multidisciplinary intensive pulmonary rehabilitation (PR) program between 2021 and 2024 were screened for inclusion.

Inclusion criteria were as follows: (1) confirmed diagnosis of COPD according to the GOLD recommendations [8]; (2) initiation of treatment with FF/UMEC/VI at the first visit; (3) age between 40 and 80 years; (4) no acute exacerbation within four weeks prior to enrollment; (5) availability of at least two records approximately 12.0 ± 1.0 months apart, namely T0 (enrollment) and T12 (follow-up).

Exclusion criteria included patients already receiving inhaled corticosteroids, those with concomitant severe cardiovascular disease, ongoing malignancies, history of lung transplantation or major lung surgery, inability to perform spirometry, and patients with incomplete or missing relevant data.

Clinical, laboratory, and functional data were extracted from medical records corresponding to the predefined timepoints (T0 and T12).

The study was conducted in accordance with the Declaration of Helsinki of the World Medical Association and reported following the STROBE guidelines [9]. Since this was a retrospective analysis of prospectively collected data under a different protocol approval number (Comitato Etico Campania 2 n° 3/20 Maugeri del Registro), the approval of the local Institutional Review Board was waived.

### 2.2 Study Procedures and Outcomes

The primary endpoint was complete clinical stability (CS) at one year, defined by the simultaneous presence at T12 of the following three criteria: (1) absence of acute exacerbations between T0 and T12; (2) reduction of at least 2 points in the CAT score; (3) decline in FEV□ ≤ 100 mL between T0 and T12.

All patients underwent at least one cycle of multidisciplinary intensive PR between T0 and T12.

### 2.3 Statistical Analysis

Continuous variables are presented as mean ± standard deviation (SD) or median (interquartile range), depending on distribution normality. For normally distributed variables, a two-sided Student’s t-test was applied for between-group comparisons. For non-normally distributed variables, the Mann–Whitney U test was employed. Changes between T0 and T12 were analyzed using paired Student’s t-test or Wilcoxon signed-rank test, as appropriate. Post-hoc sensitivity analyses were conducted to estimate statistical power. Potential predictors of clinical stability were evaluated using logistic regression models.

## 3. Results

### 3.1 Study population

Forty-seven consecutive individuals for whom all outcomes were available were enrolled in the study (Table 1). Of them, 38 (80.9%) were males, the mean age was 73.6 ± 5.2 years, and all had a relevant smoking history, with a mean packs/year of 19.2±5.4. Cardiovascular comorbidities were reported in the 72.7% of all cases. The exacerbation rate at baseline was moderately high, with a median of 2.0 (2.0-3.0) events/year, while a high disease burden was testified by the mean CAT score values of 27.0 ± 5.6 points. Lung function was overall impaired, with a mean FEV_1_ of 1.04 ±

**Table 1.** Characteristics of the enrolled population and comparison between T0 and T12. Significant comparisons have been highlighted in bold. Continuous variables are expressed as means ± standard deviation, unless otherwise specified. At the post-hoc power analysis, 1-β was always ≥ 0.900. Abbreviations: BMI, body mass index; mMRC, modified Medical Research Council scale for dyspnea; CAT, COPD assessment test; FEV_1_, forced exhaled volume in the first second; 6MWD, six-minute walking distance; BEC, blood eosinophil count.

| Variable | T0<br>(N = 47) | T12<br>(N = 47) | Significance |
| --- | --- | --- | --- |
| Age, years | 73.6±5.2 | - | - |
| BMI, kg/m<sup>2</sup> | 24.7±5.5 | 24.2±4.9 | 0.683 |
| Males, n (%) | 38 (80.9%) | - | - |
| Smoking history, n (%) | 47 (100) | - | - |
| Exacerbation rate | 2.0 (2.0 – 3.0)* | 1.0 (0 – 1.0)* | **<0.001** |
| mMRC score | 2.9±0.5 | 2.8±0.8 | 0.429 |
| CAT score | 27.0±5.6 | 24.9±5.0 | **<0.001** |
| FEV<sub>1</sub>, L | 1.04±0.41 | 0.97±0.48 | 0.053 |
| FEV<sub>1</sub>, % | 42.4±15.4 | 41.4±19.4 | 0.659 |
| 6MWD**, m | 140.0 (103.8 – 232.8)* | 210.0 (140.3 – 289.0)* | **<0.001** |
| BEC<sup>†</sup>, cell/mm<sup>3</sup> | 240.0 (185.0 – 340.0)* | 145.0 (99.5 – 295.0)* | **0.023** |

\* Data are expressed as median (interquartile range).
\*\*6MWD was available in 24 out of 47 patients.
<sup>†</sup>BEC was available in 22 out of 47 patients.

0.41 L, corresponding to a percent predicted FEV_1_ of 42.4 ± 15.4 %. In 24 out of 47 (51.1%) subjects, the 6MWD was available both at T0 and at T12; at baseline, the median unadjusted 6MWD was 140.0 (IQR: 103.8 – 232.8) m.

### 3.2 Effects of FF/UMEC/VI on clinical parameters at T12

At T12, a significant drop in the exacerbation rate was observed, with a decrease of 64.5%; similarly, a significant positive variation was observed in the CAT score (P < 0.001; 1-β: 0.904), as shown in Table 1. While FEV_1_ variations did not achieve statistical significance, 6MWD significantly raised to a median of 210.0 (IQR: 140.3 – 289.0), P < 0.001. *vs*. baseline values (1-β: 0.956).

### 3.3 Clinical Stability at T12

A total of ten patients (21.3%) met the three criteria for CS at T12, while the number of patients achieving at least one criterion varied significantly, as shown in Figure 1.

**Figure 1.**
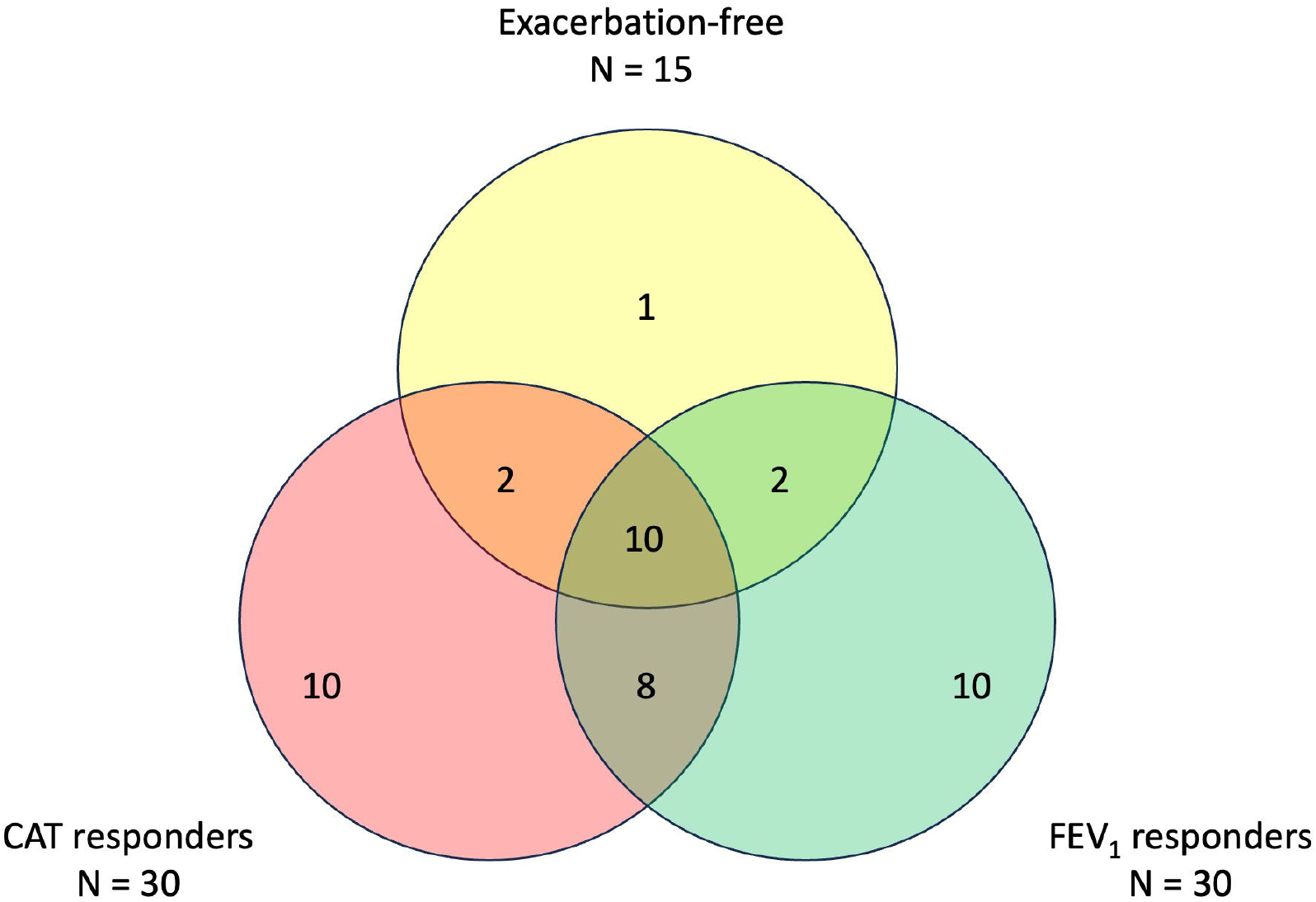
Overlap of responders for CAT, FEV_1_ and exacerbations at T12. Collectively, 15 (31.9%) individuals were exacerbation-free, 30 (63.8%) subjects were considered as sole CAT responders, while 12 (25.5%) individuals responded in terms of exacerbations and CAT score; similarly, 30 (63.8%) participants were FEV_1_ responders and 12 of them responded also in terms of exacerbations. Finally, 18 (38.3%) individuals were both CAT- and FEV_1_-responders. Abbreviations: CAT, COPD Assessment Test. FEV_1_, forced exhaled volume in the first second.

A comparison in the baseline data and variations in key outcomes between individuals reaching CS and those who did not (non-CS) has been presented in table 2. At baseline, a significant difference was found in the exacerbation rate [2.0 (IQR:2.0 – 2.0) among CS *vs*. 3.0 (IQR:2.0 – 3.0) among non-CS, P = 0.020 (1-β: 0.997)]. The two groups also differed in the mMRC scores, which were significantly lower in the cCS group (Table 2). Finally, a numerically but not statistically significant difference was found in lung function parameters, particularly FEV_1_, which was higher in the CS group (P = 0.076). Interestingly, a statistically significant (albeit tendential) difference was observed in the variation (Δ) in the 6MWD between T12 and T0 in the two groups. In fact, individuals in the cCS group presented with a mean increase in the 6MWD of 70.8±45.8 m, while in non-CS individuals the observed variation was 37.9±27.1 (P = 0.048, 1-β: 0.917).

**Table 2.** Comparison between participants who achieved complete Clinical Stability (CS) and those who did not. Continuous variables are expressed as means ± standard deviation, unless otherwise specified. At the post-hoc power analysis, 1-β was always ≥ 0.900. Abbreviations: mMRC, modified Medical Research Council scale for dyspnea; CAT, COPD assessment test; FEV_1_, forced exhaled volume in the first second; 6MWD, six-minute walking distance; BEC, blood eosinophil count.

| Variable | CS (N = 10) | Non-CS (N = 37) | Significance |
| --- | --- | --- | --- |
| Age, years | 76.3±4.8 | 72.9±5.1 | 0.068 |
| Males, n (%) | 10 (100) | 28 (75.7) | 0.083 |
| BMI, kg/m<sup>2</sup> | 26.8±5.5 | 24.1±5.5 | 0.263 |
| Smoking history, n (%) | 10 (100) | 10 (100) | >0.999 |
| **T0** |  |  |  |
| Exacerbation rate | 2.0 (2.0 – 2.0)* | 3.0 (2.0 – 3.0)* | **0.020** |
| mMRC score | 2.5±0.5 | 3.0±0.5 | **0.017** |
| CAT score | 25.8±5.2 | 27.4±5.8 | 0.438 |
| FEV<sub>1</sub>, L | 1.24±0.45 | 0.98±0.39 | 0.076 |
| FEV<sub>1</sub>, % | 50.1±12.0 | 40.3±15.7 | 0.074 |
| 6MWD**, m | 210.0(130.5 – 309.5)* | 140.0 (100.0-220.0)* | 0.183 |
| BEC<sup>†</sup>, cell/mm<sup>3</sup> | 235.0 (142.5 – 785.0)* | 240.0 (190.0 – 340.0)* | 0.965 |
| **T12** |  |  |  |
| Exacerbation rate | 0 (0 – 0)* | 1.0 (1.0-1.0)* | **<0.001** |
| mMRC score | 2.9±0.9 | 2.8±0.8 | 0.611 |
| CAT score | 22.0±4.6 | 25.7±4.9 | **0.036** |
| FEV<sub>1</sub>, L | 1.41±0.52 | 0.86±0.40 | **<0.001** |
| FEV<sub>1</sub>, % | 58.9±17.0 | 37.1±17.6 | **0.002** |
| 6MWD**, m | 302.0 (216.0 – 345.0)* | 190.0 (122.0 – 242.0)* | 0.053 |
| Δ6MWD, m | 70.8±45.8 | 37.9±27.1 | **0.048** |
| BEC<sup>†</sup>, cell/mm<sup>3</sup> | 110.0 (89.5 – 675.0)* | 160.0 (100.0 – 240.0)* | 0.940 |

\* Data are expressed as median (interquartile range).
\*\*6MWD was available in 24 out of 47 patients (5 CS, 19 non-CS).
<sup>†</sup>BEC was available in 22 out of 47 patients (5 CS, 17 non-CS).

### 3.4 Potential predictors of cCS

Based on univariate analysis, a multivariate logistic regression model was constructed to explore potential predictors of cCS. After adjusting for age and sex, neither baseline mMRC score nor baseline exacerbation rate was independently associated with a higher likelihood of achieving CS (OR 0.231, 95% CI 0.037–1.424 and OR 0.112, 95% CI 0.011–1.153, respectively). However, in a model including age, sex, and baseline CAT score, the baseline exacerbation rate was a strong predictor of CS (OR = 0.082, 95% CI 0.009–0.754). This indicates that, independent of age, sex, and symptom burden, each additional exacerbation per year was associated with a 92% decrease in the odds of achieving CS after 12 months.

## 4. Discussion

Our study highlights that continuous treatment with the triple inhaled therapy FF/UMEC/VI significantly reduces the exacerbation rate and improves health-related quality of life (HRQoL) in individuals with COPD. Importantly, this is the first study to document the prevalence of CS, defined by the absence of exacerbations, reduction in symptom burden (CAT score), and preserved lung function, in a real-life COPD cohort treated with FF/UMEC/VI. After one year of therapy, a notable 21.3% of patients achieved CS, highlighting the potential of triple therapy to induce substantial and clinically meaningful disease control. Additionally, we observed a trend toward better functional improvement, as measured by the 6MWD in those patients who reached CS, thus suggesting that clinical stability may translate into tangible benefits in exercise capacity.

The ability of FF/UMEC/VI to reduce exacerbation frequency and improve HRQoL is well supported by robust evidence from randomized controlled trials, such as the IMPACT trial [10], as well as real-world observational studies [11] . Our findings align with these reports and somewhat extend them by quantifying the proportion of patients who achieve a multidimensional state of clinical stability over a prolonged treatment period in a real-life setting.

Acute exacerbations have been shown to negatively impact disease progression, morbidity, and mortality in COPD [12] and, therefore, achieving CS is of paramount importance and for routine clinical practice. In this clinical setting, preventing exacerbations and maintaining symptom control and lung function represent key therapeutic goals. The CS construct, which integrates these dimensions, provides a comprehensive measure of treatment success beyond isolated clinical endpoints. Our study’s demonstration that a sizeable subset of patients can reach this state with FF/UMEC/VI treatment underscores the therapy’s potential to alter disease trajectory and improve long-term outcomes.

Comparisons with other studies reveal some variability in the reported rates of clinical stability, likely reflecting differences in cohort characteristics, definitions of clinical stability, and treatment regimens. In this context, ELLITHE was a multicenter, open-label, non-interventional effectiveness study conducted in Germany from 2020 to 2022 [13]. In the subsequent post-hoc analysis [7], the same study group demonstrated that a substantial proportion of patients with COPD experienced clinically meaningful benefits from once-daily FF/UMEC/VI therapy across multiple dimensions of disease control at 12 months. Over half of the cohort (53.3%) showed significant improvement in symptoms, as indicated by a reduction of at least two points in the CAT score, while 36.7% achieved a clinically relevant increase in lung function (≥100□mL FEV□). Notably, the majority of patients (90.2%) remained free from moderate or severe exacerbations during the study period. Despite these individual improvements, only about one-fifth (22.1%) of patients met responder criteria across all three outcomes simultaneously. This value was confirmed in our retrospective analysis of a real-life COPD subjects’ cohort, but striking differences emerge when comparing the single items composing the CS definition, particularly regarding the exacerbation-free status, which was reached in the 31.9% of our study population. This discrepancy may derive from the difficulty of assessing exacerbations in real-life setting, probably deriving from misdiagnosis (e.g. COPD titration vs. mild/moderate exacerbation), poor inhaler technique and other potential confounding factors, such as heart failure [14].

A clinically mindful finding of our study is the observed trend toward improved 6MWD in patients achieving CS. This evidence, albeit with the relevant limitations described below, paves the way for novel studies aimed at investigating a potential role of cCS in the field of PR. It is in fact known that exercise capacity is a critical determinant of daily functioning, independence, and prognosis in COPD [15], while improvements in 6MWD have been linked to reduced hospitalization risk and mortality [16]. Given the evidence that patients who reached CS also tended to improve their exercise tolerance, it might be hypothesized that stable disease control may facilitate greater physical activity and rehabilitation gains, potentially creating a positive feedback loop that further enhances outcomes. This putative additive effect of FF/UMEC/VI therapy on top of PR also suggests that pharmacologic optimization can potentiate rehabilitation benefits by reducing exacerbations and improving baseline symptom control. Also, a reduction in lung inflammation induced by pharmacological treatment might reflect onto systemic aspects, such as endothelial dysfunction, which has been recently acknowledged as a key feature of COPD-associated cardiovascular risk [17].

In line with previous reports [18], we observed that patients reaching CS were burdened by a lower exacerbation rate at baseline and presented with a trend for improved baseline spirometric values, thus hinting at the possibility that a precocious introduction into treatment of FF/UMEC/VI in eligible patients might be desirable in order to increase the odds of stabilizing their condition.

However, it is important to acknowledge relevant limitations that affect the generalizability of our findings. Our cohort consisted exclusively of patients engaged in a PR program, which might have confounded the effects on HRQoL, lung function, and exacerbations. While PR effects are typically temporary, the interplay between optimized pharmacotherapy and rehabilitation requires further study to delineate their respective contributions and optimize integrated care pathways. The relatively small sample size may have limited statistical power to detect differences and fully characterize the prevalence of CS in the broader COPD population. Additionally, a selection bias may be identified when considering that only patient records with complete clinical data were included for evaluation; furthermore, data collection during the SARS-CoV-2 pandemic may have influenced outcomes due to changes in healthcare access, patient behavior, and infection risk.

## 5. Conclusion

Our study adds relevant real-world evidence supporting the use of triple inhaled therapy with FF/UMEC/VI to reduce exacerbations, improve quality of life, and promote clinical stability in COPD. The association between CS and enhanced exercise capacity suggests that achieving and maintaining stability should be a key therapeutic target, not only to reduce acute events but also to maximize functional status and patient-centered outcomes.

Future research should focus on larger, prospective studies to validate our findings and explore the mechanisms by which pharmacologic therapy and pulmonary rehabilitation interact. Investigating the longitudinal impact of achieving CS on disease progression, healthcare utilization, and survival would also provide valuable insights. Moreover, evaluating strategies to sustain PR benefits in conjunction with optimized triple therapy may help refine integrated COPD management approaches.

## Data Availability

All data produced in the present study are available upon reasonable request to the authors

## Notes

### Competing Interest Statement

The authors have declared no competing interest.

### Funding Statement

This study was partially funded by the Ricerca Corrente funding scheme of the Italian Ministry of Health

### Author Declarations

The requirement of ethical approval was waived by Comitato Etico Territoriale Campania 2 for the studies involving humans because the study is a retrospective analysis of prospectively collected data. The studies were conducted in accordance with the local legislation and institutional requirements. The ethics committee/institutional review board also waived the requirement of written informed consent for participation from the participants or the participants' legal guardians/next of kin because a written informed consent had been already obtained for the original prospective trial (approval number 3/20 Maugeri del Registro).

